# PYCR1 Levels Track with Premature and Chronological Skin Aging

**DOI:** 10.1101/2023.05.24.23289766

**Authors:** Kortessa Sotiropoulou, Saniye Yumlu, Tomoko Hirano, Michael Maier, Abigail Loh, Peh Fern Ong, Onn Siong Yim, Chunping Liu, Emmanuel Vial, Umut Altunoğlu, Sheela Nampoothiri, Deepthi de Silva, Björn Fischer-Zirnsak, Hülya Kayserili, Poh San Lai, Oliver Dreesen, Kenji Kabashima, Uwe Kornak, Nathalie Escande-Beillard, Bruno Reversade

## Abstract

De Barsy syndrome is a recessive progeroid disease classified under the group of cutis laxa syndromes. The disease is attributed to loss-of-function mutations in *PYCR1* or *ALDH18A1*, leading to premature skin aging. Here we report five *PYCR1* pathogenic alleles and a mouse knockout model of the disease. Through these investigations, we have confirmed the key role of PYCR1 in preventing dermal thinning and other connective tissue abnormalities. However, it remains unknown whether endogenous PYCR1 levels undergo changes during normal aging. To address this query, we examined its levels in cultured human cutaneous fibroblasts subjected to induced or replicative senescence. In both instances, PYCR1 levels dropped and correlated with the loss of proliferative capacity. Furthermore, we validated the relevance of these findings *in vivo*, by comparing young and chronologically aged human skin, and found that the levels of PYCR1 in the dermis, but not the epidermis, significantly decreased with age. Our results confirm that the loss of PYCR1 is a driver of human skin aging and that its levels in healthy individuals can serve as a biomarker for connective tissues undergoing normal chronological aging.

## MAIN

De Barsy Syndrome (DBS, <u>MIM:614438, MIM:219150</u> and <u>MIM:616603</u>) is a recessive progeroid disease belonging to cutis laxa syndromes, which are characterized by wrinkled, thin and translucent skin that is more prominent at the hands, feet and the abdomen (Zampatti et al. 2012). Patients’ skin exhibits fragmentation and rarefaction of elastic fibers and only mild collagen irregularities (Reversade et al. 2009). The syndrome is caused by loss-of-function mutations affecting either PYCR1 *(pyroline-5-carboxylate reductase 1)* or P5CS (*pyroline-5-carboxylate synthase)*, the two main enzymes responsible for proline anabolism (Figure 1A) (Fischer-Zirnsak et al. 2015; Reversade et al. 2009).

**Figure 1:**
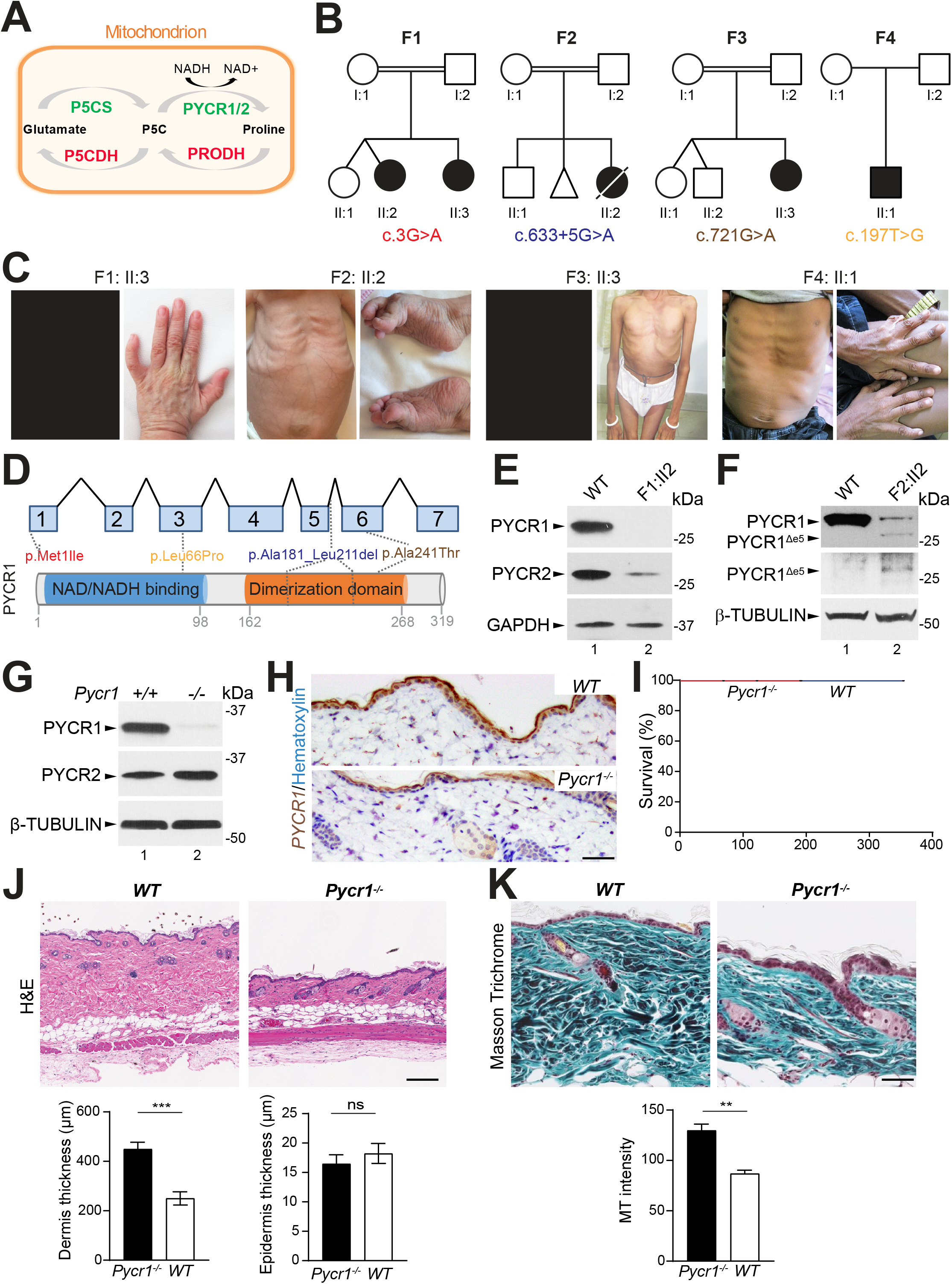
PYCR1 is required to maintain skin integrity in humans and mice. **(A)** Schematic representation of the mitochondrial proline metabolic pathway. P5CS mediates the conversion of glutamate into Δ1-pyrroline-5-carboxylate (P5C), which is then reduced into L-proline by PYCR1 or PYCR2 in a NAD(P)H dependent manner. The reverse reaction is catalyzed by PRODH and P5CDH. **(B)** Pedigrees of affected individuals from three consanguineous families and one non consanguineous family segregating with autosomal recessive cutis laxa type II. Square, male; circle, female; black shading, affected proband; double lines, consanguineous marriages; diagonal line, deceased. The patients in family 1 carry a homozygous change c.3G>A in the start codon of *PYCR1*. Family 2 carries an intronic single base change c.633+5G. The patients in families 3 and 4 carry missense mutations: c.721G>A and c.197T>G respectively. **(C)** Pictures from affected individuals from 2 to 13 years old show the typical progeroid triangular face (F1-II:3), thin and translucent skin in the abdomen (F2-II:2, F4-II:1), wrinkled skin affecting primarily the hands and feet (F1-II:3, F2-II:2, F4-II:1), prognathism and growth retardation (F3-II:3). **(D)** Schematic view of the PYCR1 gene and protein monomer indicating that p.Met1Ile and p.Leu66Pro are located in the NAD/NADH binding domain, whereas the p.Ala181_Leu211del and p.Ala241Thr are located in the dimerization domain. **(E)** Western blot showing that c.3G>A leads to a PYCR1 protein-null allele with concomitant reduction of PYCR2 protein levels. **(F)** Western blot showing that the p.Ala181_Leu211del mutation leads to a significant loss of full length PYCR1 protein (top band, lane 2), and the formation of smaller isoform PYCR1^Δe5^ detected by a commercial antibody directed against residues 1-171 of PYCR1 (bottom band, lane 2) and a custom antibody specific to PYCR1^Δe5^ (bottom band, lane 2). β-tubulin protein serves as a loading control. **(G)** Western blot of primary dermal fibroblasts shows that PYCR1 is absent in *Pycr1*^*-/-*^ mice compared to WT cells. PYCR2 level is not affected by deletion of PYCR1. β-tubulin serves as a loading control. **(H)** Immunohistochemistry staining with PYCR1 (brown) antibody and hematoxylin (blue) illustrates that there is no *PYCR1* expression in the dermis of the dorsal skin of *Pycr1*^*-/-*^animals at 80 days. Signal observed in the epidermis is background. Scale bar, 100 μm **(I)** Kaplan-Meier survival curves over 50 weeks show normal longevity of *Pycr1*^*-/-*^ (n = 16 males, red line) compared to *WT* (n = 8 males, blue line) littermates. **(J)** Hematoxylin and Eosin (H&E) staining illustrates that the dermis of *Pycr1*^*-/-*^ mice at day 80 is visibly thinner compared to that of *WT*, whereas the epidermis appears unchanged. Quantification shows that the dermis thickness of *Pycr1*^-/-^ compared to *WT* mice is significantly reduced in the dorsal skin. No significant change in epidermal thickness can be documented. Scale bar, 100 μm. Lengths were calculated by averaging three measurements in each dorsal section (n = 6 mice/genotype). Two-tailed Student’s t test, ns p>0.05 ***p<0.001. **(K)** Masson Trichrome staining shows reduced number of collagen fibers (green) in *Pycr1*^-/-^ mice (muscles are stained red and nuclei black). Quantification confirms that the number of collagen fibers is significantly reduced in *Pycr1*^-/-^ compared to *WT* mice. Scale bar, 100 μm. Stainings were analyzed by measuring intensity using ImageJ. Images from three independent skin samples were analyzed and averaged in each group. Two-tailed Student’s t test, **p<0.01.

Unexpectedly, proline levels in sera or primary fibroblasts from affected PYCR-deficient individuals are within normal range arguing against proline auxotrophy (Dimopoulou et al. 2013; Escande-Beillard et al. 2020; Fischer-Zirnsak et al. 2015; Reversade et al. 2009). The exact molecular etiology of PYCR1-deficiency remains largely enigmatic and its link to normal chronological aging has not been examined.

We report here five additional patients (Figure 1B) that displayed prominent acrogeria and a typical DBS face gestalt (Figure 1C). Sanger sequencing revealed four novel germline mutations in *PYCR1* that were homozygous in the affected children and had never been described in UK10K or gnomAD databases. In family 1, a single homozygous c.3G>A variant leading to the loss of p.Met1 resulted in the total absence of endogenous PYCR1 and a significant reduction in PYCR2 level (Figures 1A, D and E). In family 2, a homozygous intronic splice site mutation c.633+5G>A, which caused an in-frame exon 5 skipping, resulted in reduced full-length PYCR1 and a smaller isoform corresponding to PYCR1^Δe5^ (Figures 1B, D and F). Patients from families 3 and 4 segregated the following germline recessive variants c.721G>A; p.(Ala241Thr) and c.197T>C; p.(Leu66Pro), respectively (Figures 1B and D). These five patients extend the allelic spectrum of DBS and confirm the importance of PYCR1 for ensuring skin homeostasis in humans.

To obtain a potential animal model of DBS, we generated a constitutive *Pycr1* mouse knockout line. mRNA and protein analysis showed that *Pycr1* mutant mice had little, to no, residual *Pycr1* left, consistent with a complete deletion of both alleles (Figures 1G, H and Supplementary Figures S1A-D). *Pycr1*^*-/-*^ mice were born to Mendelian ratios, were viable and fertile with a lifespan comparable to *WT* siblings (Figure 1I and Supplementary Figure S1E).

We next sought to characterize the effects of *Pycr1* depletion in the dermal lineage, which is the most severely affected tissue in DBS patients. While their epidermis remained unaffected, *Pycr1*^*-/-*^ mice exhibited significant dermis thinning compared to *WT* (Figure 1J), with significantly less collagen fibers (Figure 1K and Supplementary Figures S1F-G).

Although the thinning of the dermis seen in *Pycr1*^*-/-*^ animals suggests a partial resemblance to the human disease, the mice’s overall phenotype remains relatively normal. We thus speculate that PYCR2 might have the potential to compensate for the loss of PYCR1 in mice. In *Pycr1*^*-/-*^ mouse fibroblasts, the levels of PYCR2 remained unchanged (Figure 1G). In contrast, human fibroblasts lacking PYCR1 see a knock-off effect resulting in a significant decrease in PYCR2 levels (Figure 1E). This species-specific difference, where the loss of *Pycr1* can be rescued by *Pycr2* in mice but not in human cutaneous fibroblasts, suggest that only double *Pycr1/Pycr2* knockout mice could phenocopy the full spectrum of DBS symptoms.

The progeroid phenotype caused by the loss of *PYCR1* in humans and its validation in mice highlight the importance of PYCR1 for sustaining the integrity of the skin. It is, however, unknown whether endogenous PYCR1 levels correlate with natural aging. To address this, we examined proline metabolism in the context of cellular senescence. The overexpression of *P16, TP53* or *BRAF*^*V600E*^ is known to trigger stress-induced senescence (van Deursen 2014). Human dermal fibroblasts were thus engineered to overexpress these transgenes. Cellular senescence was assessed by Lamin B1 (LB1) and DCR2, two inversely regulated senescence markers (Althubiti et al. 2014; Dreesen et al. 2013), and by the loss of proliferation measured by PCNA. The RNA level of proline metabolic pathway components was unchanged (Supplementary Figure S2A), but the protein levels of endogenous PYCR1 and PYCR2 were decreased, while the levels of P5CS, PRODH and PYCR3 remained stable (Figure 2A). The changes in PYCR1 levels were specific to senescence, and not a general consequence of growth arrest, as contact inhibition induced-quiescence did not alter PYCR1 protein levels (Supplementary Figure S2B).

**Figure 2:**
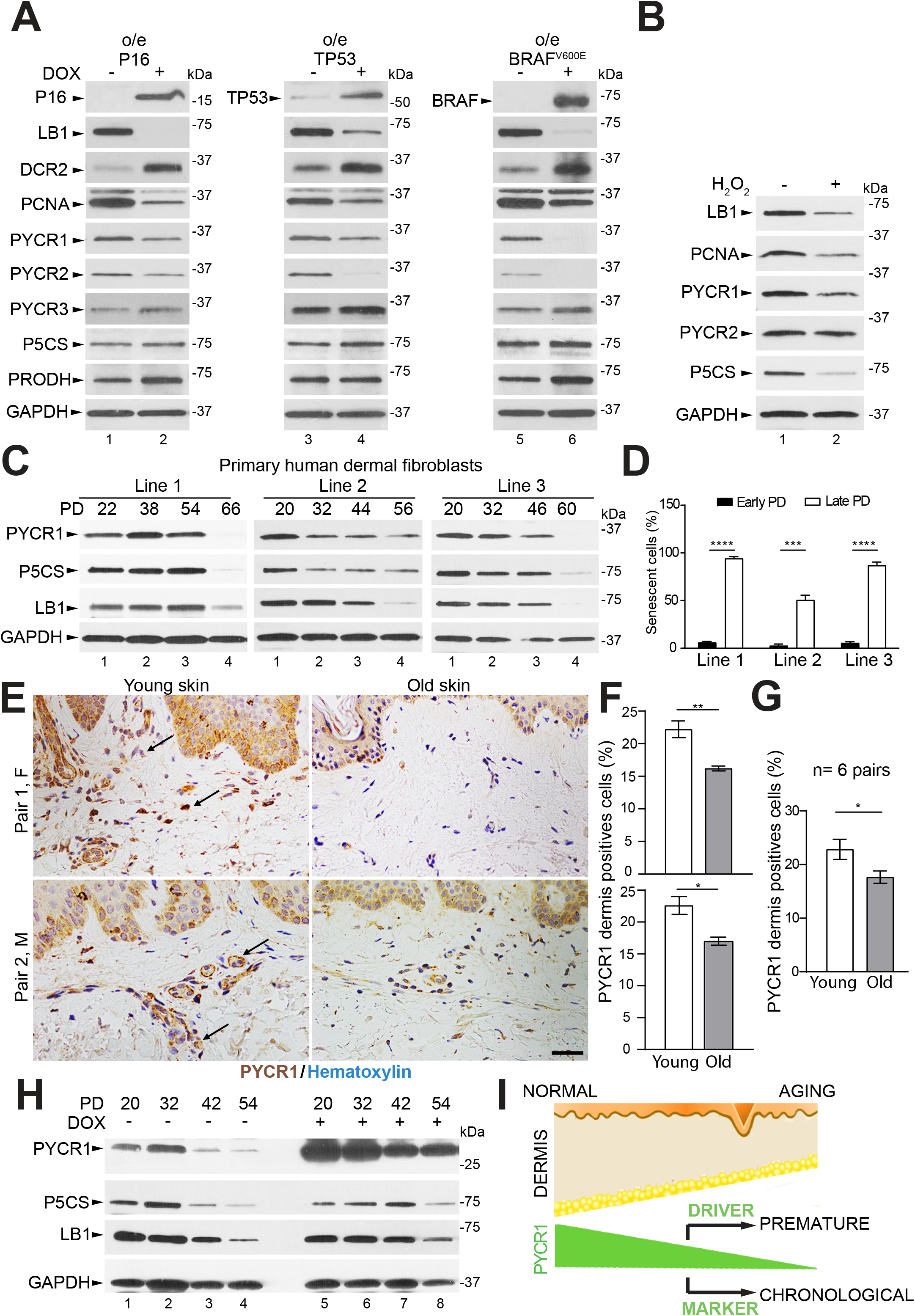
PYCR1 levels are reduced in human senescent fibroblasts and aged dermis. **(A)** Western blot data show overexpression (o/e) of the transgenes *P16, TP53* and *BRAF*^*V600E*^ upon 4 days of doxycycline treatment. Levels of LB1 and DCR2 which are established senescence markers, decreased and increased respectively. PCNA, an established proliferation marker, is reduced upon doxycycline treatment. Endogenous PYCR1 and PYCR2 protein levels are reduced while PYCR3, P5CS and PRODH do not change significantly. **(B)** Senescence-induced by oxidative stress (H_2_O_2_) leads to LB1, PCNA, PYCR1 and P5CS protein level reduction, and no changes in PYCR2 levels. GAPDH serves as loading control. **(C)** Three independent primary dermal fibroblast lines derived from three young healthy individuals were serially cultured to replicative senescence. Western blots on the three dermal primary fibroblast lines show that endogenous PYCR1, P5CS and LB1 protein levels decrease with loss of replicative capacity in human primary fibroblasts. GAPDH levels are unchanged and serve as a loading control. **(D)** β-galactosidase staining quantification shows that the percentage of blue stained fibroblasts in the late PDs is significantly higher compared to the early PDs. Error bars indicate mean ± SEM. Two-tailed Student’s t test ***p<0.001, ****p<0.0001 (n = 6). **(E)** Representative immunohistochemistry images illustrating PYCR1 expression (brown) and hematoxylin (blue) on skin samples from young (15 to 30 years old) and old donors (70 to 80 years old). Black arrows indicate PYCR1 positive staining in the dermis. Pairwise comparison was performed according to gender (M, male; F, female) and anatomical region of the skin where the biopsy was taken. Scale bar, 50 μm. **(F)** Staining quantification for pairs 1 and 2 reveals that PYCR1 level is lower in the dermis of old compared to young individuals (n=5 pictures/pair). Percentages of PYCR1 positive cells were calculated in 5 independent pictures from each sample. In each picture 3 sections of 20 cells were measured and averaged. Error bars indicate mean ± SEM. Two-tailed Student’s t test, *p<0.05, **p<0.01, ***p<0.001, ****p<0.0001. **(G)** Graph demonstrates that the combined dermis quantification of PYCR1 positively stained cells is significantly lower in old compared to young individuals (n = 6 pairs). Error bars indicate mean ± SEM. Two-tailed Student’s t test, *p<0.05. **(H)** Western blot illustrates that PYCR1 and P5CS protein levels reduce in *WT* cells upon replicative senescence, like LB1, whose reduction indicates that senescence is induced. After overexpression of PYCR1, LB1 level reduces, suggestive of no changes in senescence. GAPDH serves as loading control. **(I)** Schematic representation summarizing the findings of this study. Congenital loss of PYCR1 results in premature aging, while normal aging leads to PYCR1 reduction in the dermal lineage.

The reduction of PYCR1 protein level was also observed upon oxidative stress, while PYCR2 protein levels remained stable and those of P5CS were reduced (Figure 2B). Although stress-induced senescence occurs physiologically, chronological telomere erosion serves as a stable contributing factor to normal aging. Thus, we sought to further characterize the effect of replicative senescence by culturing three healthy human fibroblast lines for extended population doublings (PD). Increased SA-β-galactosidase staining and reduced LB1 protein level (Figures 2C-D and Supplementary Figure S2C) confirmed the occurrence of senescence, and were correlated with a gradual decline of PYCR1 and P5CS protein levels particularly at late time points (Figure 2C). Collectively, these findings along with the induced senescence experiments (Figure 2A), confirm a positive correlation between senescence and reduction of endogenous PYCR1 protein levels in human dermal fibroblasts.

To validate these results *in vivo*, we obtained healthy skin tissue from six young (aged 15-30 years) and six older (aged 65-80 years) individuals. Immunohistochemistry revealed that endogenous PYCR1 expression was significantly reduced in the dermis, but not epidermis, of old compared to young individuals (Figures 2E-G and Supplementary Figure 2D-E). These results are consistent with PYCR1 being an essential component for the homeostasis of dermal lineages, which is acutely compromised in DBS patients (Zampatti et al. 2012).

Overall our data suggests that increasing PYCR1 activity in humans could potentially delay or mitigate connective tissue aging. To assess this hypothesis, human dermal fibroblasts engineered to overexpress PYCR1 were cultured to replicative senescence. In this *in vitro* model, senescence assessed by the level of LB1 could not be significantly rescued by overexpression of PYCR1 (Figure 2H). However, several studies have shown that overexpression of PYCR1 is linked to hyperproliferation in a variety of solid tumors (Chen et al. 2019; Kuo et al. 2020; Loayza-Puch et al. 2016; Yan et al. 2019), suggesting that greater-then-normal PYCR1 expression can promote proliferation *in vivo*. Having been first documented in hepatocellular carcinoma, this phenomenon is likely to be lineage restricted and may become mobilized only once a cell has been oncogenically transformed. At this stage, it is fair to state that the genetic loss of PYCR1 is a driver of premature skin aging, while it can also serve as a marker of normal chronological aging in the human dermis (Figure 2I). It thus remains to be demonstrated if rewiring a cell’s metabolic flux towards increased proline anabolism, via PYCR1/2 and P5CS gain of activity, can mitigate the effect of chronological cellular aging.

## MATERIALS AND METHODS

### Subjects and informed consent

All participants or their parents/guardians in this study provided written informed consent. The clinical diagnosis of De Barsy syndrome was confirmed in all patients by experienced clinicians in Turkey, India and Sri Lanka. Skin biopsies were obtained from individuals from family 1 and family 2. Peripheral blood samples were collected from individuals I:1, I:2, II:2 and II:3 from family 1, I:1, I:2 and II:2 from family 2, and I:1, I:2 and II:2 from family 3, I:1, I:2 and II:3 from family 4 and I:1, I:2 and II:1 from family 5. Genomic DNA extraction was performed using standard methods. All human studies were reviewed and approved by the Turkish, Japanese and Singaporean (A*STAR IRB 2019-087) institutional review boards. All patients gave their consent to publish their photographs.

### Sanger sequencing

Segregation of *PYCR1* (NM_006907.2) variants was examined by bidirectional Sanger sequencing in genomic DNA samples from each family member and patients available. PCR primers were designed as shown in Table 1.

### Generation of primary skin cells

Primary skin cells were isolated from fresh skin biopsies. Biopsies were decontaminated using a cocktail of antibiotics. Keratinocytes were isolated from the epidermis using 0.25% trypsin and grown in keratinocyte media with a 3T3 layer, which had been mitotically inactivated (Rheinwald and Beckett, 1981). The dermis was used to obtain primary dermal fibroblasts. Fibroblasts were isolated using 1X collagenase D (Roche, 11088882001).

### Cell culture

Human dermal primary fibroblasts were cultured in DMEM high glucose (HyClone) supplemented with 10% FBS (HyClone), 1% penicillin / streptomycin (Gibco) and 1% L-glutamine (Gibco) in 37°C and 5% CO_2_. Human dermal primary keratinocytes were cultured in keratinocyte serum free media (KSFM) in 37°C and 5% CO_2_.

### Immunoblot analysis

Whole cell lysates of human dermal primary fibroblasts were isolated with cOmplete™ Lysis-M EDTA-free mammalian cell protein extraction reagent kit (Merck) completed with 1x protease inhibitors (Merck), 10% SDS (1^st^-Base) and 0.1M DTT, followed by electrophoresis in 4-20% pre-cast SDS polyacrylamide gels (BIO-RAD). Immunoblot analysis was performed using PVDF membranes probed with the following antibodies: rabbit anti-PYCR1 (Proteintech) 1:2000, rabbit anti-PYCR2 (Proteintech) 1/500, rabbit anti-PYCR3 (Abnova) 1/500, rabbit anti-P5CS (Novus) 1/500, goat anti-P5CS (Abnova) 1/500, rabbit anti-PRODH (Abcam) 1/500, mouse anti-p16 (Biolegend) 1/500, mouse anti-p53 (Santa Cruz) 1/500, mouse anti-V5 (Santa Cruz) 1/5000, rabbit anti-DCR2 (Abcam) 1/1000, rabbit anti-p21 (Santa Cruz) 1/500, rabbit anti-p27 (Santa Cruz) 1/100, mouse monoclonal anti-Lamin B1 (hybridoma clone BBLB1 c7) 1/20, rabbit anti-PCNA (Abcam) 1 μg/ml, mouse anti-GAPDH (Santa Cruz) 1/2000.

### Mice

The Pycr1 knockout line was generated by homologous recombination in murine ES cells. A targeting vector with 4.3 kb and 3.3 kb homology regions to the *Pycr1* locus was constructed to flank exons 3 to 6 with loxP sites in the same orientation to obtain an excision of the genomic region between the two loxP sites after cre-recombination (Supplementary Figure S1a). ES cells were screened by Southern blot and aggregated with tetraploid morulae. For genotyping gDNA was extracted from tail cuts of 21-25 days old mice using lysis buffer with the following components: 5 mM EDTA, 100 mM Tris pH8, 200 mM NaCl and 0.2% SDS. Nested PCR was performed to amplify the targeted regions of *Pycr1* (Supplementary data Figure S1b), followed by 1% agarose gel electrophoresis. PCR primers used are depicted in Table 1. Dorsal skin was isolated from *Pycr1* knockout and wildtype mice, fixed in 4% paraformaldehyde, embedded in paraffin and sectioned.

C57Bl/6 mice were bred and maintained under standard laboratory conditions 12 hours light/dark cycle and had free access to food and water. All mouse procedures were done in compliance with IACUC protocols #171263 under the approval of the Biological Resource Center, A*STAR and Agri-Food & Veterinary Authority of Singapore.

### Quantitative PCR

RNA isolation from transformed dermal fibroblasts was performed with RNeasy Mini Kit (Qiagen). 1 microgram of RNA was reverse transcribed with iScript (BIO-RAD) and subsequently quantitative PCR was performed with SYBR green (Thermofisher Scientific) and the primers depicted in Table 1.

### Immunohistochemistry

Human skin biopsies of 2-4 mm diameter were obtained from affected unaffected individuals and fixed immediately with 10% neutral buffered formalin (Sigma-Aldrich, #HT501128) then embedded in paraffin and sectioned into 5 µm slices. Deparaffinization, ethanol rehydration, epitope retrieval and immunostaining with PYCR1antibody (13108-1-AP, Proteintech) were automatically performed by using the Leica Bond Auto Stain system. Mouse skin sections (5 μm) were deparaffinized and rehydrated for either Hematoxylin and Eosin, Masson’s trichrome or Herovici stainings or immunohistochemistry following the above protocol. Images were taken with Olympus BX51 upright.

### Generation of inducible-senescence human fibroblasts

The BRAF^V600E^, TP53, P16 and PYCR1 overexpressing constructs were generated by inserting the transgenes into the pTRIPZ vector carrying a doxycycline inducible promoter (Dharmacon). The *PYCR1* transgene was isolated by PCR using PYCR1-PCS2 construct as template. *BRAF*^*V600E*^ was PCR amplified from pBABEbleo-FLAG-BRAF^V600E^ (Addgene ID: 53156), *TP53* from pcDNA3-wTP53 (a gift from David Lane, A*STAR) and *P16* from pCMV-p16INK4A (Addgene ID: 10916). The resulting PCR products were digested with AgeI and MluI and ligated into pTRIPZ using the primers depicted in Table 1. Transduction into human fibroblasts was conducted according to manufacturers’ protocols (Dharmacon) as described before (Chojnowski et al. 2015).

### Senescence induction by BRAF^V600E^, TP53 and P16 overexpression

Cells were grown at 80% confluence and transgene expression was induced by addition of 1 μg/ml of doxycycline to the cell culture media. Protein and RNA samples were isolated after 4 days of transgene overexpression by doxycycline addition.

### Oxidative stress assay

Human dermal primary fibroblasts and fibroblasts overexpressing the PYCR1 transgene were treated with 300 μM of H_2_O_2_ for 30 min at 37°C. Protein samples were harvested 6 days post-treatment.

### Senescence associated-β-galactosidase staining

Human dermal primary fibroblasts were fixed in 2% PFA and 0.2% glutaraldehyde solution in PBS for 15 min at room temperature, washed with PBS, stained in complete staining solution: 5mM Potassium Ferrocyanide (Sigma), 5mM Potassium Ferricyanide (Sigma), 2 mM MgCl_2_, 150 mM NaCl, 40 mM citric acid (Sigma) and 1 mg/ml X-gal/ N,N-dimethylformamide and incubated at 37°C for 12 hours in the dark. Images were collected using light microscopy.

## Data Availability

All data produced in the present work are contained in the manuscript

## DATA AVAILABILITY STATEMENT

No datasets were generated or analyzed during the current study.

## ACKNOWLEDGEMENTS

We are grateful to all the individuals and the families for their participation and generous help in this research. We also extend a special thanks to Carine Bonnard, Alicia Yap and the Asian Skin Biobank (ASB) for providing human keratinocytes and feeders, Samsul Sapiee and the BRC mouse facility for outstanding mouse husbandry. We are grateful to all members of the Reversade laboratory for support and constructive feedback. BF-Z received funding from the Deutsche Forschungsgemeinschaft (DFG; FI 2240/1-1). U.K. received funding from the Deutsche Forschungsgemeinschaft (DFG). B.R. is an investigator of the National Research Foundation (NRF, Singapore), Branco Weiss Foundation (Switzerland) and an EMBO Young Investigator, and is supported by an inaugural Use-Inspired Basic Research Fund from the central fund from the Agency for Science & Technology and Research (A*STAR) in Singapore. N.E.B. is funded by a 2232 International Fellowship for Outstanding Researchers Program of Scientific and Technological Research Council of Turkey (TÜBİTAK) (Project No: 118C318).

## AUTHOR CONTRIBUTIONS

Conceptualization: NEB, BR; Data curation: NEB, KS; Formal analysis: NEB, KS, SY; Funding acquisition: BR; Investigation: KS, AL, SY, PFO, OSY, CL, DS, BF-Z, LP, OD; Methodology: KS, NEB, PFO, OD, SY, UK; Project administration: NEB, BR; Resources: TH, UA, SN, DS, OD, HK, UK; Supervision: NEB, BR; Writing-original draft: KS, Writing-review & editing: KS, MM, NEB, BR, BF-Z, UK, OD, KK, LPS.

