## Supplementary Table 1 for "PYCR1 Levels Track with Premature and Chronological Skin Aging"

**Supplementary Table 1:** List of primers sequences

| | |
|---|---|
| **Sanger sequencing** | |
| *PYCR1* exon 1 fw | 5'-CCCTAATCCGCTCTCGAT-3' |
| *PYCR1* exon 1 rev | 5'-CCTGCAGAAGTGGAAAGAGG-3' |
| *PYCR1* exon 5 fw | 5'-TCTGCACGGAGGTGGAAG-3' |
| *PYCR1* exon 6 rev | 5'-CCCCTAGTCCCCCTAGTGAC-3' |
| *PYCR1* exon 6 fw | 5'-CCTGGGGCTTCACTGGGA-3' |
| *PYCR1* exon 6 rev | 5'-CCCACAGTAACCCAGGGAAC-3' |
| *PYCR1* exon 6 fw | 5'-GGTGGAAGAGGACCTGATTG-3' |
| *PYCR1* exon 6 rev | 5'-ATCTGCTGAGTGCGTGAATG-3' |
| *PYCR1* exon 3 fw | 5'-CAGCATTCTCTGTGCCATTC-3' |
| *PYCR1* exon 3 rev | 5'-TTCTCCTCCTTCCCTTCTGG-3' |
| **Genotyping** | |
| Pycr1 fw | 5'-TTTTGCTGCACACCTGACTC-3' |
| Pycr1 rev1 | 5'-AGCCAGGAAGAGCACATCAC-3' |
| Pycr1 rev2 | 5'-GCCTTGGCTCTGCTTTACAC-3' |
| **Quantitative PCR** | |
| *PYCR1 fw* | 5'-TGCTCATCAACGCTGTGG-3' |
| *PYCR1 rev* | 5'-CACCTGCTCCTGGTCAGC-3' |
| *Pycr1 fw* | 5'-TGTGAAAGTTGTGGGTCGTG-3 |
| *Pycr1* rev | 5'-ACCGTAGCTTGGTCCATGTC-3' |
| PYCR2 fw | 5'-TCGGCTCACAAGATAATAGCC-3' |
| PYCR2 rev | 5'-TTCACCGTCTCCTTGTTGC-3' |
| PYCR3 fw | 5'-CCCAGACCCTGCTGGGGACG-3' |
| PYCR3 rev | 5'-CTCCACGGCGCTCATGGTGG-3' |
| ALDH18A1 fw | 5'-TCTCGTCCTGACTGTCTACCC-3' |
| ALDH18A1 rev | 5'-TAACAAGCCATTGCCACTTG-3' |
| PRODH fw | 5'-CCCTGCTTCGGCACTAGAG-3' |
| PRODH rev | 5'-GGGCCCTGGTATTGCTTGTCC-3' |
| GAPDH fw | 5'-TGAACCACCAACTGCTTAGC-3' |
| GAPDH rev | 5'-GGCATGGACTGTGGTCATGAG-3' |
| PYCR1 fw | 5'-AACCGGTCGCCACCATGAGCTGGGGC-3' |
| PYCR1 rev | 5'-ACGCGTCCTAGGCTAATCCTTGCCCGC-3' |
| **Cloning** | |
| *BRAF*$^{V600E}$ fw | 5'-TGCGCAAACCGGTGGCCACCATGGGTAAGCCTATCCCTAACCCTCTCCTCGGTCTCGATTCTACGGCGGCGCTGAGCGGTG-3' |
| *BRAF*$^{V600E}$ rev | 5'-TGCGCAAACGCGTCGTAGGTCAGTGGACAGGAAACGCACC-3' |
| *TP53* fw | 5'-TGCGCAAACCGGTGGCCACCATGGAGGAGCCGCAGTC-3' |
| *TP53* rev | 5'-TGCGCAAACGCGTCCTAGGTCAGTCTGAGTCAGGCCCTTC-3'' |
| *P16* fw | 5'-TGCGCAAACCGGTGGCCACCATGGAGCCTTCGGCTGACTGG-3' |
| *P16* rev | 5'-TGCGCAAACGCGTCCTAGGTCAATCGGGGATGTCTGAG-3' |
