## Supplemental Figure 1 for "PYCR1 Levels Track with Premature and Chronological Skin Aging"

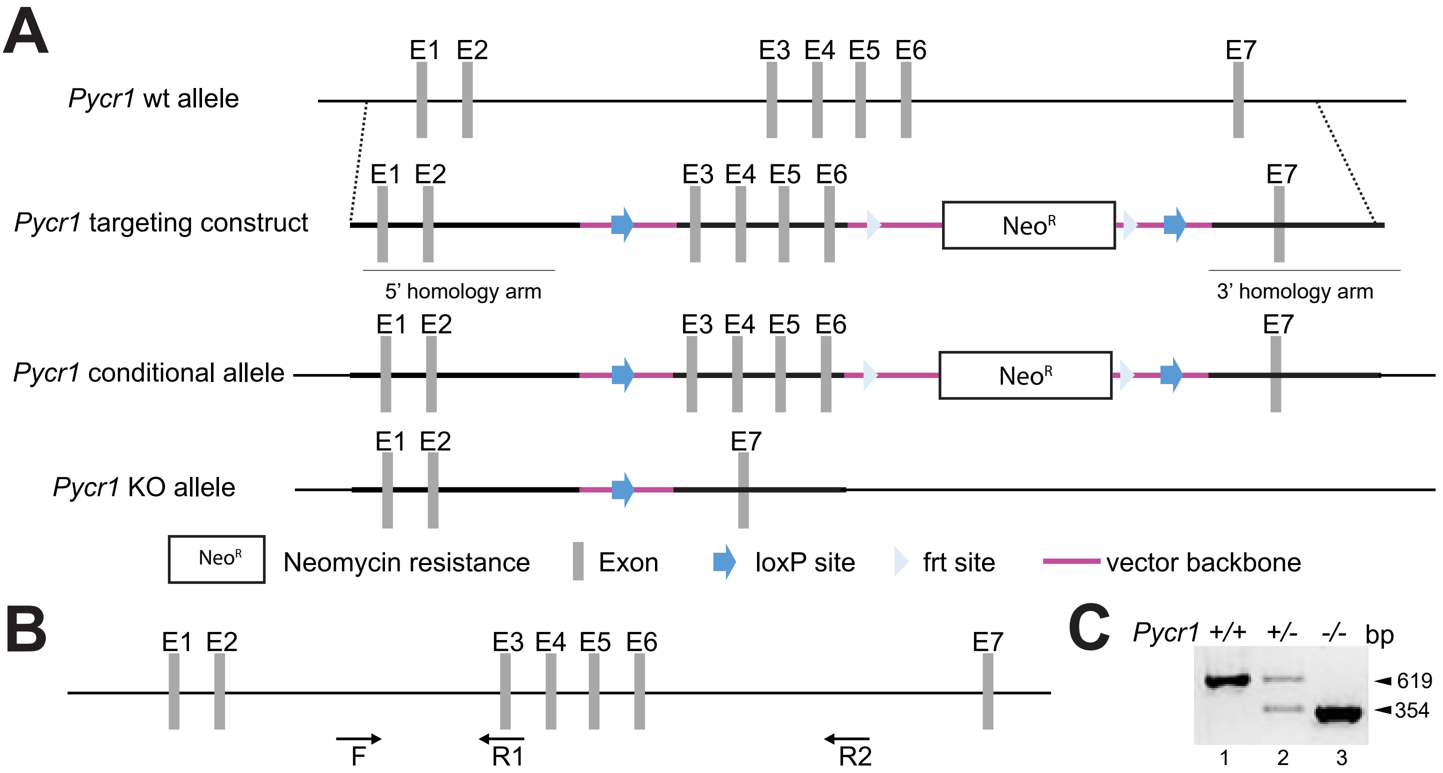


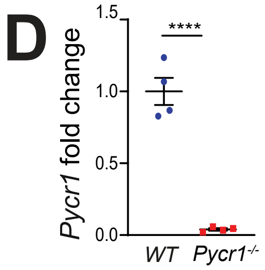


E

| Crosses | +/+ | +/- | -/- | L | p-value | %P |
|---|---|---|---|---|---|---|
| *Pycr1*$^{+/-}$ x *Pycr1*$^{+/-}$ | 21 | 50 | 35 | 12 | 0.092 $^{n.s.}$ | 34 |
| *Pycr1*$^{+/-}$ x *Pycr1*$^{-/-}$ | 0 | 43 | 53 | 12 | 0.146 $^{n.s.}$ | 54.5 |

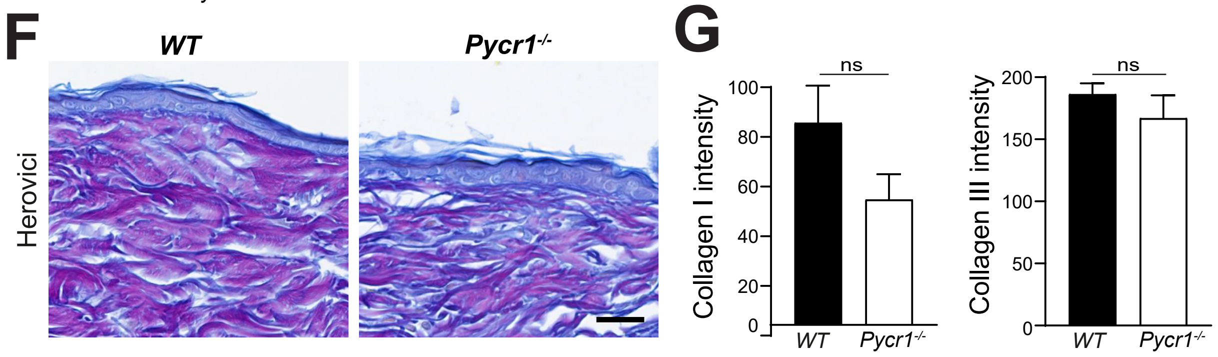


Suppl Figure 1

**Supplementary Figure S1: Generation of *Pycr1* knockout mice. (A)** Strategy and allele description for *Pycr1* knockout mice. **(B)** Schematic of gDNA from WT *Pycr1*. Arrows are depicting the primers used to genotype *Pycr1*$^{+/+}$ (619 bp, F + R1) and *Pycr1*$^{-/-}$ (354 bp, F + R2). **(C)** Genotyping of *Pycr1*$^{+/+}$, *Pycr1*$^{+/-}$ and *Pycr1*$^{-/-}$ animals by PCR. **(D)** Quantitative real time PCR demonstrating that *Pycr1*$^{-/-}$ dermal fibroblasts have virtually no *Pycr1* transcripts left compared to WT. Two-tailed Student's t test, ****$p<0.0001$ (n = 4). **(E)** Distribution of genotypes from heterozygous intercrosses and *Pycr1*$^{+/-}$ x *Pycr1*$^{-/-}$ crosses illustrate that *Pycr1*$^{-/-}$ mice are born at expected Mendelian ratios. Two-tailed Student's t test, ns $p>0.05$. **(F)** Herovici staining shows that *Pycr1*$^{-/-}$ have less collagen I fibers (pink), whereas collagen III fibers (blue) appear unchanged. Scale bar, 100 μm. **(G)** Quantification shows no significant difference in collagen I and collagen III fibers between *WT* and *Pycr1*$^{-/-}$ mice. Intensity was calculated by averaging measurements from three mice/genotypes. Error bars indicate mean ± SEM. Two-tailed Student's t test, ns $p>0.05$, **$p<0.01$, ***$p<0.001$.
