## Supplementary Figure 2 for "PYCR1 Levels Track with Premature and Chronological Skin Aging"

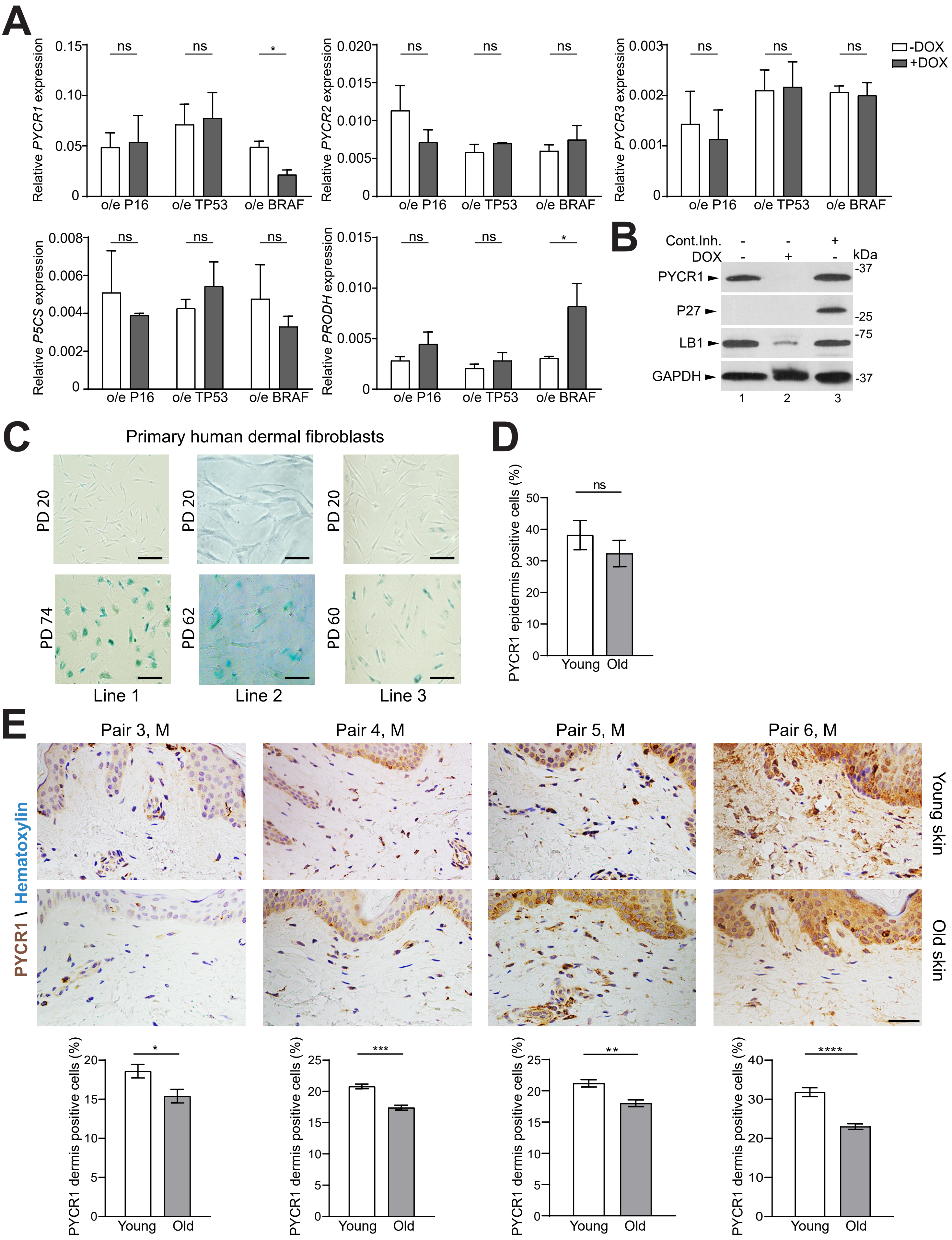


Suppl Figure 2

**Supplementary Figure S2: Induced or replicative senescence does not significantly alter the transcription of proline metabolic pathway genes.**

**(A)** Quantitative PCR data reveal that *PYCR1* mRNA levels are stable upon senescence in the *TP53*- and *P16*-overexpressing lines, but decreases in the $BRAF^{V600E}$-overexpressing line. *PYCR2*, *PYCR3*, and *P5CS* mRNA levels remain stable during senescence in the 3 cell lines, whereas *PRODH* increases upon senescence only in the $BRAF^{V600E}$-overexpressing line. n=3; Error bars indicate mean ± SEM. Two-tailed Student's t test, ns $p>0.05$, *$p<0.05$. **(B)** Western blot data from the $BRAF^{V600E}$-overexpressing line show that PYCR1, like LB1, protein level is reduced in senescence, but is stable in contact inhibition induced quiescence. The increase of P27 upon 4 days of contact inhibition confirms the quiescent state. GAPDH serves as a loading control. **(C)** Senescence-associated-β-galactosidase staining was performed at early and late population doubling (PD). Scale bar, 100 µm. **(D)** Graph illustrates that the combined epidermis quantification of PYCR1 positively stained cells is not significantly different between young and old compared individuals. Error bars indicate mean ± SEM. Two-tailed Student's t test, ns $p>0.05$. **(E)** Representative immunohistochemistry images illustrating PYCR1 expression (brown) and hematoxylin (blue) on skin samples from young (15 to 30 years old) and old donors (65 to 80 years old). Pairwise comparison was performed according to gender (M, male; F, female) and anatomical region of the skin where the biopsy was taken. Staining quantification reveals that PYCR1 level is lower in the dermis of old compared to young individuals. Percentages of PYCR1 positive cells were calculated in 5 independent pictures from each sample. In each picture 3 sections of 20 cells were measured and averaged. Scale bar, 50 µm. Error bars indicate mean ± SEM. Two-tailed Student's t test, *$p<0.05$, **$p<0.01$, ***$p<0.001$, ****$p<0.0001$.
